# High ward-level and within-sample diversity of *Klebsiella pneumoniae* on a Malawian neonatal unit revealed by single colony whole genome sequencing and post-enrichment metagenomics

**DOI:** 10.1101/2025.07.02.25330551

**Authors:** Oliver Pearse, Allan Zuza, Rebecca Lester, Helen Mangochi, Patricia Siyabu, Edith Tewesa, Thomas Edwards, Nicholas R Thomson, Nicholas A Feasey, Kondwani Kawaza, Chris Jewell, Patrick Musicha, Jennifer Cornick, Eva Heinz

**Author notes:** **Corresponding author’s email address:**.

## Abstract

*Klebsiella pneumoniae* is a frequent cause of antimicrobial resistant healthcare associated infections in neonates across sub-Saharan Africa, with multiple lineages associated with neonatal sepsis. However, the full diversity of circulating strains and key reservoirs facilitating transmission within hospitals is unknown. We investigated the population structure and within-sample diversity of *K. pneumoniae* in a Malawian neonatal unit. We recruited 94 mother-neonate pairs and collected regular stool samples, hand swabs, cot swabs and swaddling cloth samples. Additionally, we collected ward surface-swab samples and staff hand swabs weekly. To establish within sample diversity we employed a dual sequencing approach; (i) single colony picks from Extended-Spectrum Beta-Lactamase selective chromogenic agar for short-read whole genome sequencing*; and* (ii) post-enrichment metagenomics using plate sweeps from a non-selective agar. In total, we analysed 552 single-colony picks and 772 plate-sweeps from neonate, maternal and environmental samples. Comparing sequence types, surface antigens, antimicrobial resistance and virulence genes, and plasmid replicons, between sample types and sequencing approaches, we identified key advantages and limitations of post-enrichment metagenomics. Our approach revealed high diversity at both the ward and individual level, with a high proportion of the overall diversity likely due to Extended-Spectrum Beta-Lactamase negative organisms. ST15 and ST307 were found in high numbers using both methodologies, whilst ST14 was identified primarily from the non-selective post-enrichment metagenomic samples. Isolates and samples from ward surface swabs had more antimicrobial resistance genes and plasmid replicons than those isolated from human stool. This approach demonstrates the value of combining colony-based and metagenomic sequencing approaches, as a cost-effective alternative to shotgun metagenomics to study health care associated infections.

**Impact statement:** *Klebsiella pneumoniae* is an important cause of healthcare-associated infections (HAIs) in neonates, particularly in sub-Saharan Africa. While multiple strains can colonise a single person or surface, the full diversity of *K. pneumoniae* in the hospital environment is unknown. This study investigates ward-level and within-sample diversity of *K. pneumoniae* in a Malawian neonatal unit. We collected stool samples from babies and their mothers, as well as swabs from ward surfaces. To characterize bacterial diversity, we compared two approaches: (i) single-colony whole-genome sequencing of isolates selected under antimicrobial pressure and (ii) post-enrichment metagenomics, where entire microbial communities were sequenced from non-selective culture plates in a single run. We found high diversity of *K. pneumoniae*, with considerable diversity revealed by post-enrichment metagenomics. Samples frequently had multiple strains of *K. pneumoniae* within them and this varied by sample type. These data highlight the need to utilize methods that account for within-sample diversity when investigating *K. pneumoniae,* indicate that single colony WGS is inadequate and a combination of post-enrichment metagenomics and single colony WGS is preferable.

## Introduction

Neonatal sepsis is an important cause of under-5 mortality, with *Klebsiella pneumoniae* a frequent cause of neonatal healthcare-associated infection (HAI) in low-income countries such as Malawi^1, 2^. The high burden of *K. pneumoniae* HAI is often attributed to its ability to colonize the hospital environment^3^. The Chatinkha nursery in the Queen Elizabeth Central Hospital (QECH), Blantyre, Malawi is a tertiary neonatal unit that like many other neonatal units across Africa and Asia has historically had a high number of cases of *K. pneumoniae* infections^4,5^.

Bacteria colonizing hospital surfaces and patients may be important in dictating which HAI occur. The hospital is a complex environment, including different surfaces, items of equipment, staff members, patients, and their family members. We have some understanding of bacteria colonizing hospital surfaces in high-income countries; however, the healthcare environment is less well described in low- and middle-income countries (LMICs) where the burden of HAIs is high^6^ and the built hospital environment, access to disinfectants, and hospital footfall differs. Understanding bacterial colonization of the hospital environment in low-income countries could help optimize infection prevention control (IPC) strategies where resources are scarce.

*K. pneumoniae* readily acquires AMR genes and spreads rapidly; in 2020 in the Queen Elizabeth Central Hospital (QECH) Malawi, 83% of invasive isolates were resistant to ceftriaxone^5^. The main genetic determinant of ceftriaxone resistance in *K. pneumoniae* are extended-spectrum beta-lactamases (ESBLs), which hydrolyze 3^rd^-generation cephalosporins (3GC). ESBLs are globally widespread but pose a particular problem in sub-Saharan Africa, where therapeutic alternatives to 3GCs are typically limited and bacterial infections -including ESBL cases in general-^7^ are highly prevalent^8^. We know from both hospital^9^ and community^10^ studies in Malawi that rates of ESBL *K. pneumoniae* stool colonization are high but have no insight into prevalence in the hospital environment.

As is typical for an opportunistic pathogen that can survive in a variety of environments, the *K. pneumoniae* species complex is highly diverse^11^. *K*. *pneumoniae* can colonise the gastrointestinal tract and shares many similarities with other enteric bacteria such as *Escherichia coli*, including the presence of within-host diversity and competition^12,13,14^. However, the diversity of *K. pneumoniae* is far less well characterized than *E. coli*. Descriptive studies that do not account for within-sample diversity (e.g. taking one colony pick from an agar plate that might however contain phenotypically indistinguishable colonies representing different lineages in that sample are likely to be incomplete and biased^15^. Methodologies enabling a full coverage of within-sample diversity have previously been too expensive or unable to resolve within-species differences effectively. One approach would be to take several colonies from a single sample and perform whole genome sequencing on each colony^16^, but this is expensive and labour-intense, and there is still the potential to miss strains unless each colony on the plate is sequenced. Other approaches are 16S rRNA diversity or shotgun metagenomics, but these approaches are not able to distinguish organisms below genus level or are prohibitively expensive to use at scale respectively.

A novel approach has been to combine the strengths of both culture and metagenomics, by using a culture based “enrichment” of samples to support growth of target bacteria, followed by taking a plate sweep thus collecting all colonies on the plate, and performing metagenomic sequencing. This “post-enrichment metagenomics” approach allows for targeted analysis of the within-species diversity of organisms from specific samples that are at low abundance. It reduces cost, limits wasted sequencing effort, and reduces unnecessary compute and the linked CO_2_ impact^17^, compared to shotgun metagenomics on the whole sample. Bioinformatic tools such as the mSWEEP/mGEMS pipeline have now been developed that allow for analysis of this data at resolution to strain level^18, 19^.

In this study, we used two complementary approaches, ESBL selective single-colony pick short-read WGS and non-antibiotic selective post-enrichment metagenomic, to describe both the ESBL^+^ and overall diversity of *K. pneumoniae* from a Malawian neonatal unit. This comparison allowed us both to link key chromosome-plasmid combinations through single colony WGS and to assess the potential of post-enrichment metagenomics for transmission mapping in a real-world setting, and see how traits relevant for *K. pneumoniae* epidemiology, such as STs, K- and O-types, differ between the highly stringent single-colony resistance-selected method and when attempting to capture the full *K. pneumoniae* diversity via pre-enriched metagenomics without resistance selection.

## Methods

### Study location & design

This study was performed on the Chatinkha nursery in the QECH, in the Southern region of Malawi. Details of the study site are described in the Supplementary methods. It was a prospective cohort study conducted between June 2019 and April 2020. Mother-neonate dyads were recruited within 48 hours of neonatal admission onto the nursery. Neonates were admitted with a variety of health problems including sepsis. They were eligible for inclusion into the study if they were admitted to Chatinkha nursery, expected to be on the unit for greater than 24 hours and had a mother or guardian over the age of 18 years present that could be consented for enrollment in the study. Potential recruits on the ward were identified and their mothers or guardians were approached for informed written consent. Data were collected using Open Data Kit (ODK) on study tablets. Once collected, it was anonymized, only accessible to trained study team members, and stored on a password-protected server.

Study samples were either mother-neonate dyad associated or ward surface swab samples. Dyad associated samples were taken within 48 hours of admission, on day 3 of admission, and on day 7 of admission or discharge. The dyad associated samples were neonatal stool or rectal swabs, maternal stool or rectal swabs, maternal hand swabs, neonatal cot swabs or swaddling cloth (**Su**pp. Table 7; **Figure 1A**). Surface swab samples were taken on a weekly basis as per the Supplementary methods (Supp. Table 8). Stool samples were scooped into clear sterile 30ml plastic stool pots (Sterilin, Thermo Fisher Scientific, US). If a stool sample was not able to be provided, then a sterile rayon-tipped swab (Medical Wire, UK) was inserted into the rectum, rotated for 10 seconds and stored in Amies gel media for transport to the laboratory. An overview of the sampling strategy and subsequent laboratory processing is given in **Figure 1**.

**Figure 1.**
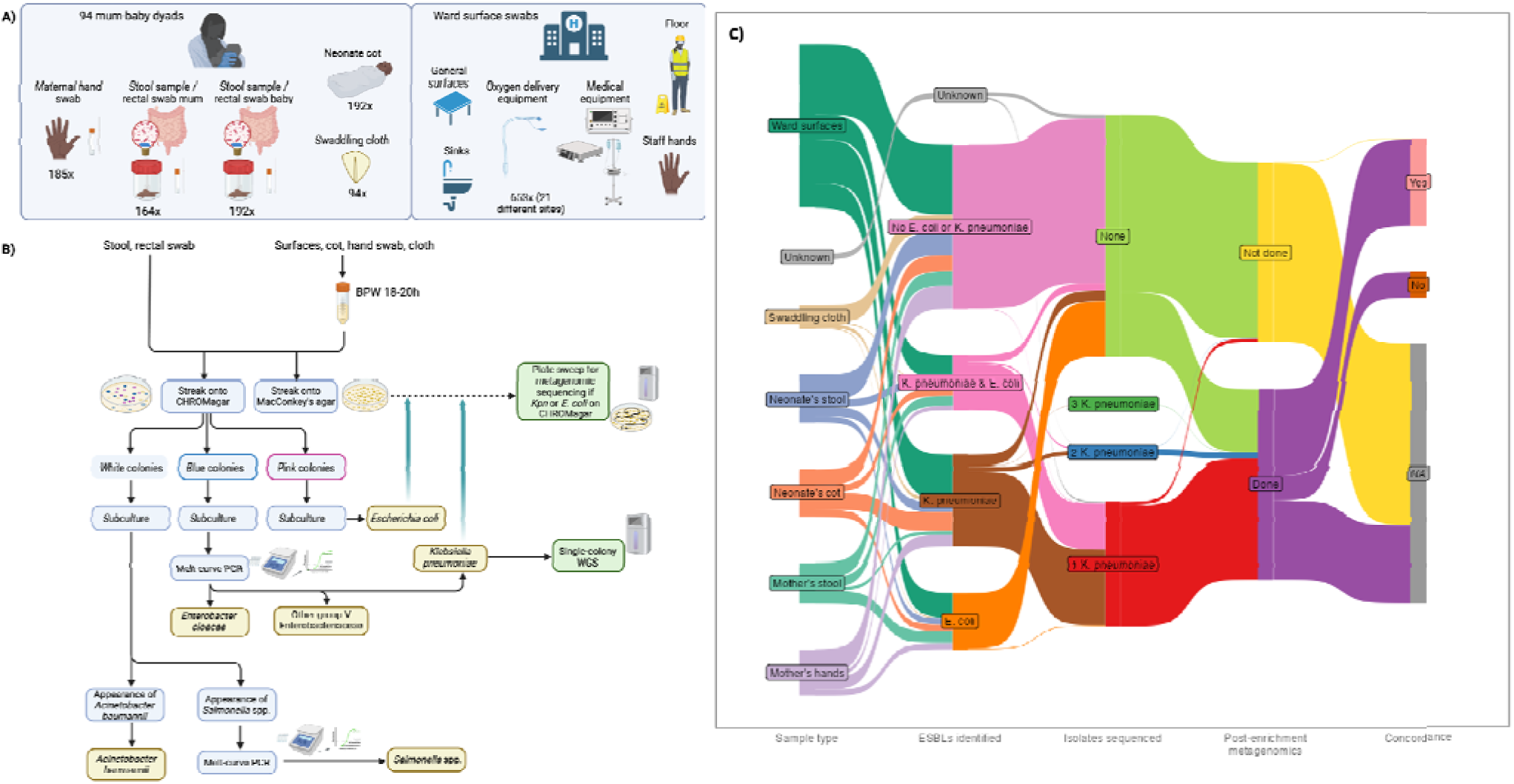
Schematic overview of **A)** the sampling strategy and **B)** the microbiological classification to select K. pneumoniae colonies for single-clone WGS or plate sweeps for metagenomic analysis. **C)** A Sankey plot showing the flow of samples. The first stage represents the different sample types, the second stage indicates whether each sample was positive for ESBL E. coli or K. pneumoniae, the third stage indicates the number of single colony WGS K. pneumoniae isolates that were analysed in this study for each sample, with red representing samples where only one K. pneumoniae was analysed, blue representing samples where two were analysed and dark green representing samples where three were analysed, the fourth stage indicates which samples underwent post-enrichment metagenomics and the fifth stage indicates whether there was concordance between the single colony WGS and the post- enrichment metagenomics.

### Laboratory procedures

Culture was undertaken at the Malawi Liverpool Wellcome Programme (MLW), on the QECH site. Ward surface swabs, cot swabs, hand swabs and swaddling cloth swabs were incubated in buffered peptone water for 18-20 hours, stool samples and rectal swabs were processed directly. Both sample types were then plated on MacConkey’s agar and ESBL selective chromogenic agar (CHROMagar, France) (**Figure 1B**). Isolates were identified based on their appearance on CHROMagar and PCR^20^ (**Supp. methods**).

A pick of a single colony of each identified isolate was stored at -80°C (single colony) in skimmed milk, tryptone, glucose and glycerine (STGG) medium. A plate sweep was taken from each MacConkey plate originating from a sample that resulted in growth of either *E. coli*, *K. pneumoniae* or both on the selective medium with a sterile swab, picking cells from all colonies on the plate, mixed and stored at -80°C in STGG medium.

### Whole genome sequencing

A total of 625 isolates identified as *K. pneumoniae* as described above were selected for sequencing, including 38 from plates with different morphologies of *K. pneumoniae* colonies, where one of each morphology was sampled. Frozen single-colony isolates were recovered from storage and purity plated on MacConkey’s agar before a single pick from the plate was selected for incubation in buffered peptone water (BPW) for 20 hours. Plate sweeps associated with samples that were positive for ESBL^+^ *K. pneumoniae*, *E. coli* or both were recovered from storage and revived by incubation in BPW for 4 hours prior to DNA extraction. The shorter incubation time for plate sweep recovery was chosen to give enough time to revive the bacteria but not enough time to allow significant competition between strains^21^. DNA was extracted either manually using QIAamp DNA Mini kits (Qiagen, Germany) or the QIAsymphony DSP Virus/Pathogen kits on the QIAsymphony automated system according to the manufacturer’s instructions (Qiagen, Germany), using on-board lysis. DNA amounts of less than 200ng were repeated. Samples were shipped to Wellcome Sanger Institute (WSI) for whole genome sequencing (WGS). At WSI, isolates underwent library preparation according to the Illumina protocol and were sequenced on the Novaseq SP using 150bp paired-end reads at 364 plex. Post-enrichment metagenomic samples underwent library preparation according to the Illumina protocol and were sequenced on the HiSeq 4000 using 150bp paired end reads at 96 plex, generating approximately 3.6 million reads per sample.

### Bioinformatic analysis

Single-colony pick reads were assembled and annotated using the automated pipeline described previously^22^ (**Supp. methods**). The identity of single colony WGS was then confirmed using Kraken v1.1.1^23^ and assembly statistics checked using CheckM v1.2.2^24^. Any sample with greater than 5% read content other than *K. pneumoniae*, *K. variicola* or Unclassified was excluded, as well as samples with <20 or >150 contigs, genome size of <5MB or >6.5MB, or greater than 5% heterozygous SNPs. Kleborate v3.0.0^25^ was used to define ST, O-loci and K-loci and virulence genes in the single colony WGS and post-enrichment metagenomic samples (**Supp. Table 2 & 5)**. ARIBA v2.14.6^26^ was used with the CARD^27^, plasmidfinder^28^ and VFDB^29^ databases to identify AMR genes, plasmid replicons and virulence factors in the isolates and post-enrichment metagenomic samples. (**Supp. Table 2 & 3**)

For single colony WGS isolates Snippy v4.6.0^30^ was used to align the reads to the *K. pneumoniae* reference genome (accession number CP000647.1)^31^ and generate SNPs. Gubbins v3.2.1^32^ was used to exclude the hypervariable regions (representing recombination and phages) of the genome. Snp-sites v 2.5.1^33^ was used to determine the constant regions for input to Iqtree as invariant sites. Iqtree v1.6.10 was used to construct a phylogenetic tree^34^, with the Modelfinder option^35^ and 10000 ultra-fast bootstraps^36^. The optimal model as determined by Modelfinder was a general time reversible model with unequal rates and unequal and empirical base frequencies, allowing for invariable sites and a discrete Gamma model^37^ with 4 rate categories (GTR + F + I + G4).

For the post-enrichment metagenomic samples the mSWEEP/mGEMS pipeline was used^18, 19^. First, a custom reference database was created, based on the standard reference database used in the original publication (Supplementary methods). This database was indexed using Themisto v0.2.0. After generation of this indexed database, raw reads from the samples were pseudo-aligned to this (**Supp. Table 2** for accession numbers), and mSWEEP v1.4.0 was subsequently used to determine the relative abundances of *K. pneumoniae* STs in the post-enrichment metagenomic samples.

*K. pneumoniae* pseudo-alignments that were at greater than 10% relative abundance in the sample were selected for further processing. mGEMs v1.0.0 was run on the raw post-enrichment metagenomic reads. Bins were assembled using Shovill v1.1.0^38^, and the quality of the assembly was checked using Checkm v1.2.2^24^. Pseudo-assemblies with a total genome length not between 5 and 6.5MB were excluded, as were pseudo-assemblies with greater than 1000 contigs. Kleborate v3.0.0 was then run on these pseudo-assemblies (**Supp. Table 5**). Further QC was undertaken by checking for concordance between the predicted ST of mGEMS and ST called by Kleborate, with pseudo-assemblies being excluded if there was discordance.

## Statistical analysis

Statistical analysis was performed in R v4.3.3^39^. The following packages were used in the analysis; here^40^, tidyverse^41^, lubridate^42^, ggplot^43^, ggpubr^44^ and ggtree^45^.

Statistics were summarized using the mean for normal data and the median for non-normal data. When comparing the difference between two means the Student’s T-test was used. The chi-squared distribution with continuity correction was used to compare the proportions of categorical variables, with Bonferroni adjustment when multiple comparisons were being made. ANOVA was used for comparison of more than two means (number of AMR genes, number of plasmid replicons)^46^.

## Ethical approvals

Ethical approval for the study was granted by the local ethics board COMREC (study number P.10/18/2499) and LSTM REC (study number 19-018). LSTM acted as the study sponsor.

## Results

### Sample overview

A total of 1483 samples were inoculated on both selective plates for ESBL enrichment, and non-selective plates for plate sweeps (**Figure 1**). Of the selective plates, 587/1483 (39.6%) were positive for ESBL *K. pneumoniae* (there was no growth of ESBL *K. pneumoniae* on 896 plates), with a total of 665 *K. pneumoniae* isolated from study samples (some samples had multiple *K. pneumoniae* colony morphologies isolated). Of all 1483 samples, 192 (12.9%) were neonatal stool samples, and 37.5% of these were ESBL *K. pneumoniae* positive. The only other sample types which were more likely to be positive for ESBL *K. pneumoniae* were the cots and the ward surface swabs (52.6% and 51.8%, respectively). Of 665 positive isolates, 625 were able to be recovered and submitted for whole genome sequencing, and after QC and de-duplication 552 isolates were taken forward for analysis, representing 525 samples.

To select the samples for post-enrichment metagenomics, we included both samples that showed growth of ESBL *K. pneumoniae* (587) and of ESBL *E. coli* (185) (**Figure 2A**). The non-selective plates derived from inoculation with these 772 samples were prepared using a plate sweep, capturing all colonies on the respective plate (methods for details). The only sample type where most non-selective samples did not contain *K. pneumoniae* was the maternal stool (Figure 2A).

**Figure 2.**
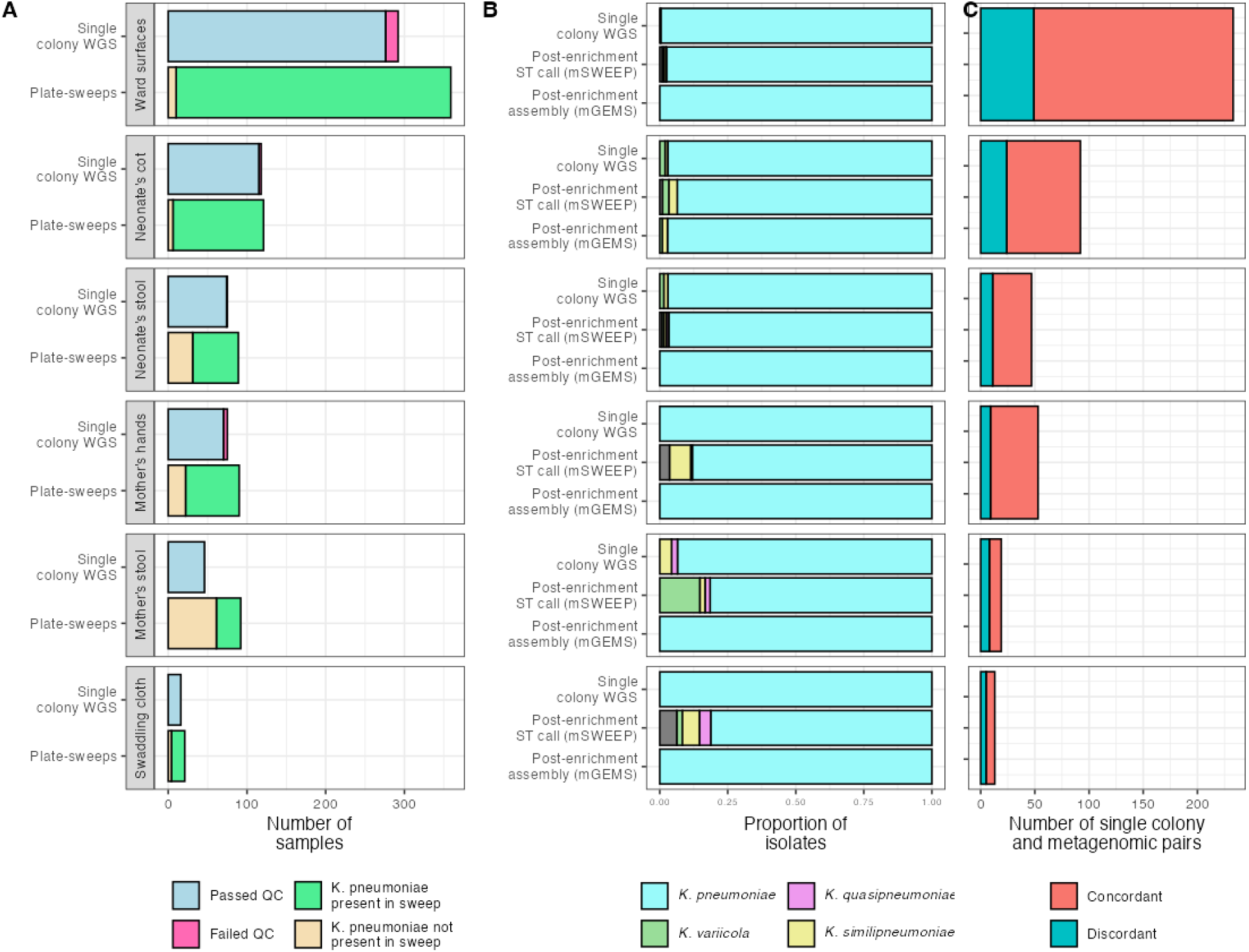
A) A bar chart showing the number of single colony WGS and post-enrichment metagenomic samples analysed during the study, separated by sample type. For the single colony WGS isolates the colours of the bars represents whether they passed QC or not. For the post-enrichment metagenomic samples the colours of the bars represents whether *K. pneumoniae* was identified in the sample using mSWEEP. **B)** A bar chart representing the proportion of the different *K. pneumoniae* subspecies that were identified using single colony WGS and in the post-enrichment metagenomic samples. *K. pneumoniae* represents *K. pneumoniae* subsp. *pneumoniae, K. variicola* represents *K. variicola* subsp. *variicola, K. quasipneumoniae* represents *K. quasipneumoniae* subsp. *quasipneumoniae* and *K. similipneumoniae* represents *K. quasipneumoniae* subsp. *similipneumoniae.* **C)** A bar chart showing the number of single colony/post-enrichment metagenomic pairs for each sample type with the colours of the bars representing whether these pairs were concordant (the same ST was identified in both modalities) or discordant (there was no overlap between the STs identified using both modalities).

### Congruence between WGS, post-enrichment metagenomic reads, and post-enrichment metagenomic assemblies

There were 457 samples (with 1235 *K. pneumoniae* ST assignments) for which we had both *K. pneumoniae* single colony WGS from ESBL selective plates and post-enrichment metagenomic data from the non-selective plates (**Figure 1B, 2)**. Of these 351/457 (76.8%) samples had a concordant call between single pick WGS and post-enrichment metagenomics, indicating that 23.2% of ESBL^+^ *K. pneumoniae* in samples were not detected from plate sweeps. There was no difference in the proportion of samples that had multiple picks per plate sequenced and whether there was concordance between the ST call in single colony WGS and the post-enrichment metagenomics. The STs for which there was a concordant call between the single pick WGS and the post-enrichment metagenomics were found at higher relative abundance in the post-enrichment metagenomics samples than those that were not isolated in the single colony WGS (mean 0.50 vs 0.15; 95% CI 0.31 - 0.39;*p* < 0.0001; unpaired t-test), and tended to have higher number of reads assigned to all groups using mSWEEP (13.8% higher; *p* = 0.004; unpaired t-test) indicating that STs found at low relative abundance and in samples at lower read depth were less likely to be identified. There was some variation in the number of mismatches (where the ESBL^+^ *K. pneumoniae* ST could not be detected in the plate sweep of the same sample) by sample type but this was not significant.

For creation of pseudo-assemblies from plate sweeps for further analysis we selected 797 *K. pneumoniae* bins (a single ST from a single sample) derived from 551 samples, that were found at greater than 10% relative abundance in the sample (with the remaining 90% being reads of other organisms or *K. pneumoniae* STs). 626 of the created pseudo-assemblies metrics were within our QC thresholds (171 were excluded) and 583/626 (93.1%) of these pseudo-assemblies had concordant results between the ST called by mSWEEP on the reads and that called by Kleborate on the pseudo-assemblies (those that did not had a lower relative abundance in the sample than those that did [26.9% vs 52%; *p* < 0.0001; unpaired t-test]). These pseudo-assemblies were used for the further analysis of the post-enrichment metagenomic data for the O- and K-type analyses.

### K. pneumoniae subspecies

The single colony WGS data revealed 543 *K. pneumoniae* subsp. *pneumoniae* isolates with 5 *K. quasipneumoniae* subsp. *similipneumoniae*, 3 *K. variicola* subsp. *variicola* and 1 *K. quasipneumoniae* subsp. *quasipneumoniae*. Analysing the subspecies by sample type showed that *K. quasipneumoniae* subsp. *similipneumoniae* was found in the mother’s stool samples, neonate’s stool samples, cot samples and ward surface swabs, that *K. variicola* subsp. *variicola* was only found in the neonate’s stool and the cots and *K. quasipneumoniae* subsp. *quasipneumoniae* was only found in the mother’s stool samples (**Figure 2B**). Using the mSWEEP call for ST from the post-enrichment metagenomics data to identify subspecies from the mSWEEP data^47^, revealed greater numbers of the less frequent *K. pneumoniae* subspecies for some sample types (8/48 [15%] of mSWEEP calls from mother’s stool identified *K. pneumoniae* subsp. *variicola* vs none from the single-pick WGS). However, many of these could not be assembled and only neonatal cots showed non- *K. pneumoniae* subsp. *pneumoniae* pseudo-assemblies; two *K. quasipneumoniae* subsp. *similipneumoniae* and one *K. variicola* subsp. *variicola* (the *K. quasipneumoniae* subsp. *similipneumoniae* and *K. variicola* subsp. *variicola* from the metagenomic samples). All remaining 580 pseudo-assemblies from plate sweeps were *K. pneumoniae* subsp. *pneumoniae* (**Figure 2B**).

### Ward level *K. pneumoniae* ST diversity using WGS and post-enrichment metagenomics

Single colony WGS identified 75 STs in 1483 samples, whilst post-enrichment metagenomic samples using mSWEEP (i.e. not subject to assembly QC) detected 111 STs representing a total of 1783 distinct *K. pneumoniae* in 772 samples. The ST most frequently observed in the single colony WGS data was ST15 (82/552 [14.9%]) followed by ST307 (69/552 [12.5%]) (**Figure 3A**). The ST most frequently observed in the post-enrichment metagenomic data was ST14 (175/1785 [9.8%]), followed by ST15 (150/1785 [8.4%)]). ST14 being found in high frequency in the post-enrichment metagenomic data was primarily due to its high prevalence in ward surface swabs. It also appeared at low abundance (119/175 [68%] were below 10% prevalence) in the post-enrichment metagenomic data and consequently could often not be assembled.

**Figure 3.**
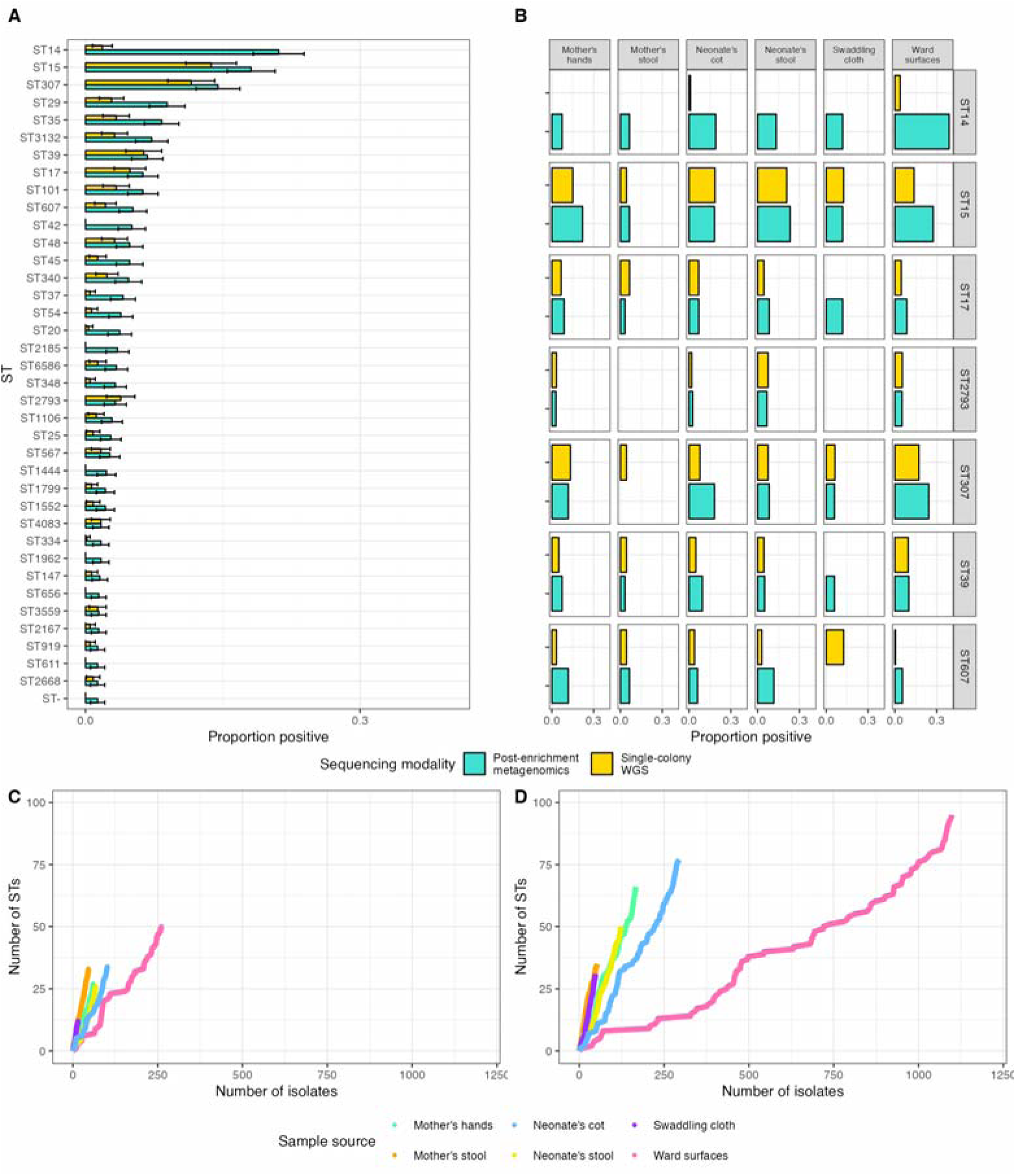
A) A bar chart showing the proportion of samples that were positive for different STs of K. pneumoniae using both single colony WGS and post-enrichment metagenomic sequencing for all sample types. **B)** A bar chart showing the proportion of samples that were positive for specific STs of K. pneumoniae by sample type, using both single colony WGS and post- enrichment metagenomic sequencing. **C)** Rarefaction curves for single colony WGS, in which isolates are randomly drawn from the overall pool of isolates in sequence, to indicate the number of new STs identified for a given number of isolates. **D)** Rarefaction curves for post- enrichment metagenomics, in which *K. pneumoniae* groupings are randomly drawn from the overall pool of groupings in sequence, to indicate the number of new STs identified for a given number of isolates.

Including only STs detected using both approaches (single-colony picks and mSWEEP analysis), four STs were found in significantly different frequencies (χ^2^ using a Bonferonni correction and a *p* threshold < 0.00076 to account for the fact that 66 comparisons were made). ST15, ST307, and ST39 were all identified more frequently in single colony WGS and ST14 in the post-enrichment metagenomics (**Supplementary Tables 2 & 4**). There were 49 STs found in the post-enrichment metagenomic samples (mSWEEP calls) not found in the single colony WGS data and 9 that were found in the single colony WGS data that were not found in the post-enrichment metagenomic samples though all were only isolated once.

In the 127 samples where no ESBL^+^ *K. pneumoniae* was identified by phenotypic processes, ST14 was identified proportionately more than in all samples combined (58/127 [46%] vs 175/638 [27.4%]; χ^2^ *p* = 0.006), implying that often ST14 are 3GC sensitive, and likely explains why it did not occur in large numbers in the single colony WGS data. No other ST was identified in significantly higher numbers. There was high diversity in these samples, with 88 STs identified in 127 samples (1.44 samples per new ST) compared to 111 STs identified in 638 samples overall (5.75 samples per new ST) implying that much of the diversity captured in post-enrichment metagenomic samples is likely due to ESBL negative isolates. In these samples there was even higher diversity in the human associated samples (maternal stool, neonatal stool, and maternal hand swabs) with 41 STs identified in 21 samples (0.51 samples per new ST).

The ST most frequently observed in neonatal stool and the cot samples in both the single colonies and post-enrichment metagenomics was ST15 (**Figure 3A**). ST14 was most frequently observed in the post-enrichment metagenomic samples from the ward surface swabs, whilst for the single colony WGS the most frequently observed ST was ST307. (**Figure 3**). Maternal stool samples had a higher-than-expected ST diversity compared to the other sample types (**Figure 3CD; rarefaction curves)**. Using bootstrapping and focusing on the single colony WGS isolates but looking at all sample types, the expected number of STs for a population of isolates the same size as the maternal stool population (46 isolates) was a median of 24 (95% CI 19 – 28), compared to 33 STs found in the maternal stool compartment. All other sample types, including pooled analysis of all human associated samples together were within the 95% CI for the number of isolates.

### Within-sample *K. pneumoniae* ST diversity using WGS and post-enrichment metagenomics

Post-enrichment metagenomic data using mSWEEP (assembly-free) revealed the median number of *K. pneumoniae* STs per sample was 2 (**Figure 4A**; Table 1) but included 54/638 (8.5%) samples with five or more STs and 14/638 (2.2%) samples with ten or more STs. There was greater ST diversity in ward surface swabs (median = 3), than maternal and neonatal stool (median = 1; **Figure 4B**). However, 23/58 (39.7.3%) of neonatal stool samples and 12/31 (38.7%) of maternal stool samples had more than one *K. pneumoniae* ST and 274/349 (78.5%) of ward surface swabs had more than one *K. pneumoniae* (**Figure 4AB**). Assessing the per-sample distribution of the major STs (**Figure 4C**) further highlights the variance in composition both based on sample type and on ST. Whilst some STs like ST35 are either completely dominant or absent, other STs (e.g. ST14, ST39) can be found at different levels of abundance within-sample. It is furthermore notable that the main STs are usually the only dominating ST, and rarely co-occur, with ST307 one of the few regularly co-occurring major STs observed, often with ST29 which was only rarely seen in single-colony WGS following ESBL selection (**Figure 3A**).

**Figure 4.**
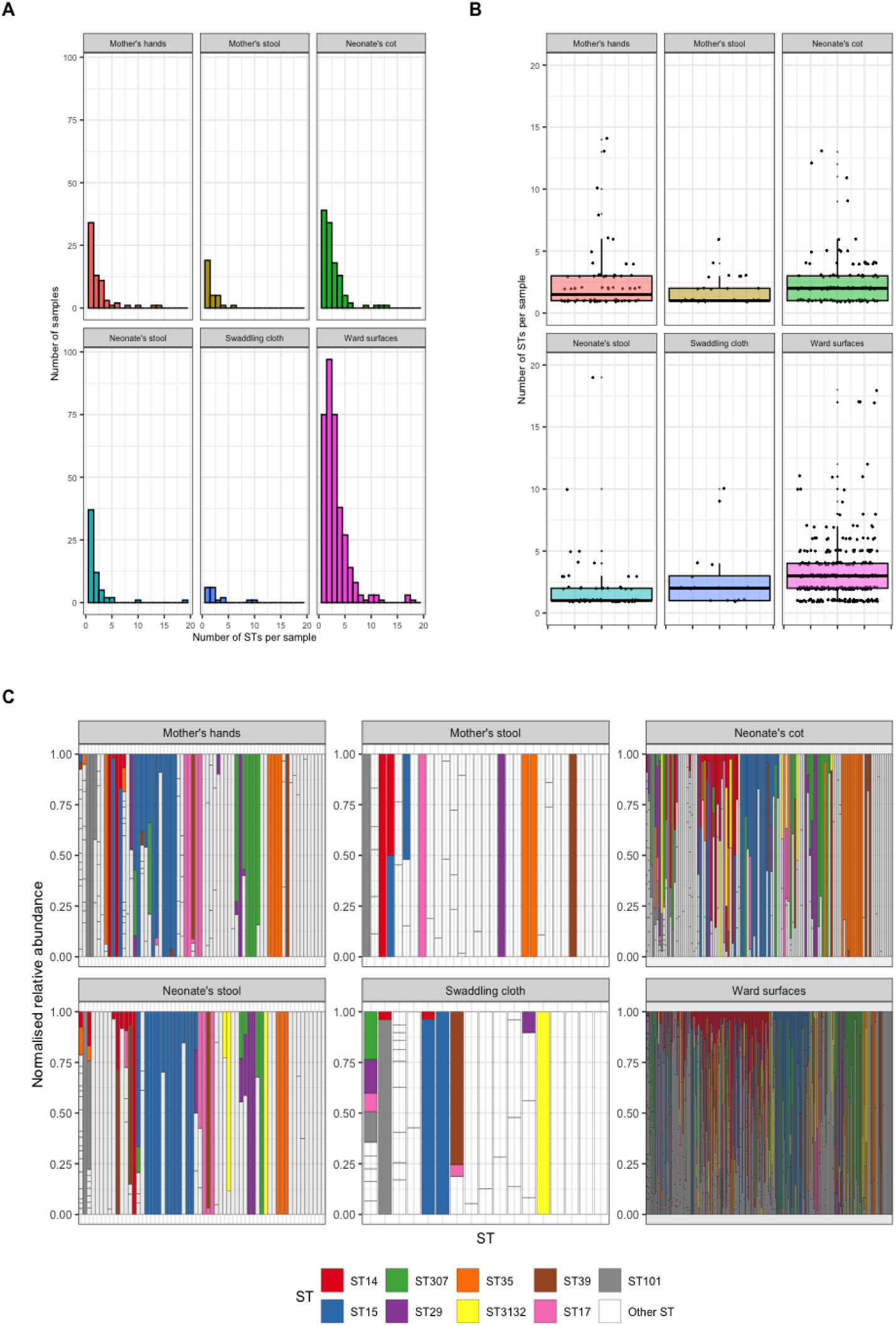
A) Distribution of *K. pneumoniae* groupings by mSWEEP for different sample types. **B)** Boxplots of *K. pneumoniae* groupings by mSWEEP for different sample tyeps. **C)** Normalised relative abundance of different *K. pneumoniae* STs per sample for different sample types. The vertical bars represent different samples and the colours represent different STs.

### Surface polysaccharide diversity

K. pneumoniae capsule (K-) and LPS O-antigen (O-type) can only be predicted from assemblies, as they depend on a set of genes present within an operon. The same nine O-types were identified using the single colony WGS data and assemblies derived from the post-enrichment metagenomic data (**Figure 5A & 7A**), with two O-types identified only using the single colony WGS data (OL104 and O12). The proportions of different O-types from the single colony data were like those in the post-enrichment metagenomic data (**Figure 5A**). The four most frequently occurring O-types in both datasets (O1/O2v1, O1/O2v2, O4 and O5) made up 513/552 (92.9%) of all isolates from the single colony WGS data and 536/573 (93.5%) of all pseudo-assemblies from the post-enrichment metagenomic data (**Figure 5A**). There was variation in the proportion of the four most frequently occurring O-loci based on sample type for single colony WGS and post-enrichment metagenomics (^2^; *p* =0.028 for single colonies; *p* = 0.04 for post-enrichment metagenomic samples) (**Figure 5B**) with neonatal cot samples having the highest proportion for the single colonies and ward surface swabs having the highest for the post-enrichment metagenomic samples. Mother’s stool had the lowest proportion for the single colonies and maternal stool the lowest for post-enrichment metagenomic samples (**Figure 5B**). O3/O3a was found only in ward surface swabs whilst O12 and OL104 were found only in mother’s stool (one isolate each).

**Figure 5.**
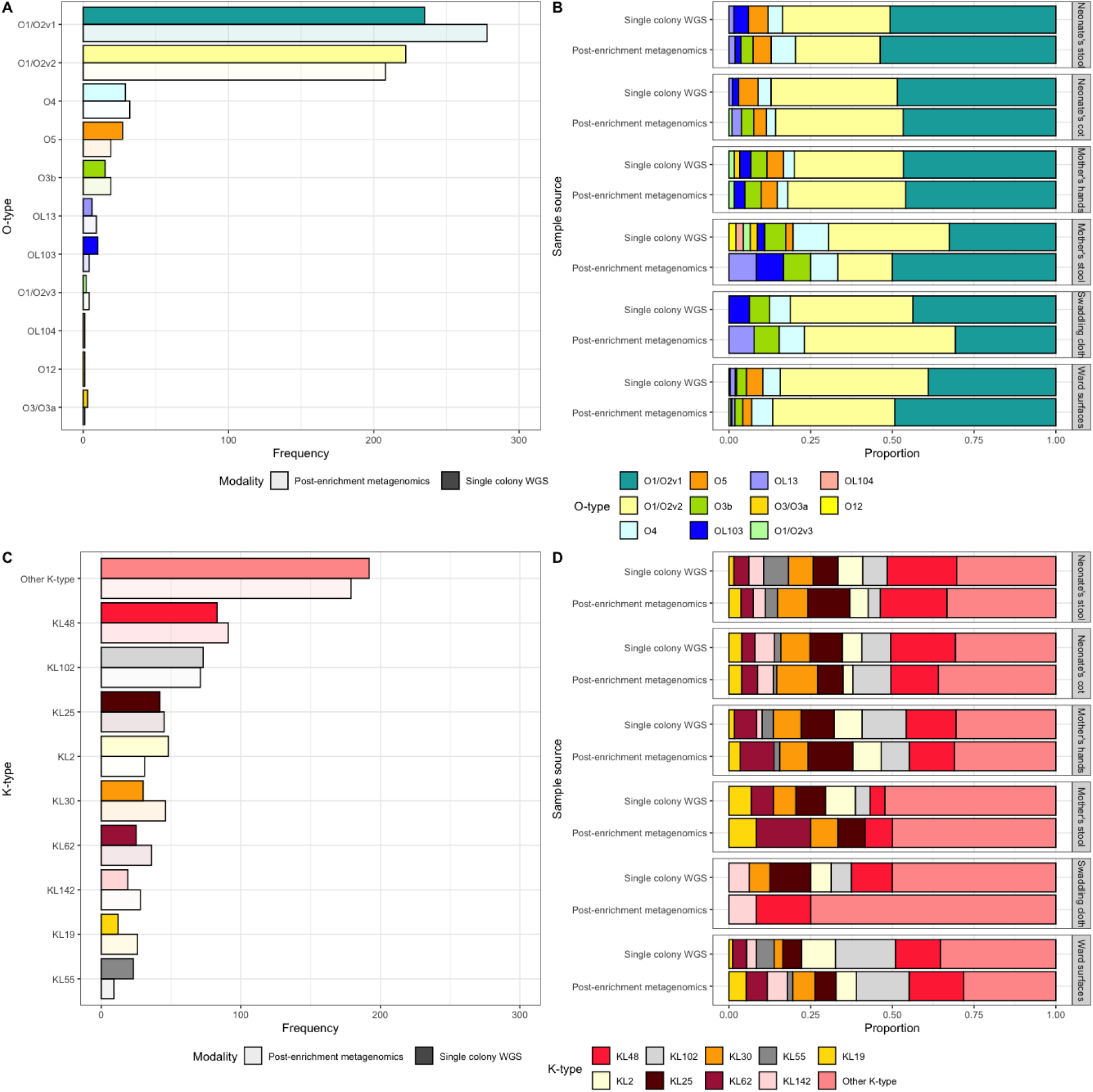
A) Bar chart showing the frequency of the different O-loci across the entire collection of samples, with the results from both single colony WGS and post-enrichment metagenomic sequencing **B)** A bar chart showing the O-loci split according to sample source, with the results from both the single colony WGS and post-enrichment metagenomic sequencing. **C)** Bar chart showing the frequency of the different K-loci across the entire collection of samples, with the results from both single colony WGS and post-enrichment metagenomic sequencing **D)** A bar chart showing the K-loci split according to sample source, with the results from both the single colony WGS and post-enrichment metagenomic sequencing.

There were 53 K-types in the single colony WGS dataset, and there were 44 in assemblies from the post-enrichment metagenomic dataset (**Figure 5C**; KL145, KL128, KL131, KL67, KL109, KL114, KL117, KL146, KL43, KL54 and KL74 were only found in the single colony data and KL22 and KL140 were only found in the post-enrichment metagenomic data). The most frequently identified K-types using single colony WGS and post-enrichment metagenomics were broadly similar (**Figure 5C**). KL48 was most frequently isolated for most sample types using both single colony WGS and post-enrichment metagenomic data. Other K-types were more frequently isolated for some sample types, such as mother’s stool (KL2 and KL25 using single colony WGS and KL62 using post-enrichment metagenomics), ward surfaces (KL102 using single colony) and swaddling cloths (KL48 and KL25 using single colony WGS; **Figure 5D**). The 13 most frequently isolated K-types using post-enrichment metagenomics represented 451/561 (80.4%) of all pseudo-assemblies and the 16 most frequently isolated K-types using single colony WGS represented 445/547 (81.4%) of all isolates (**Figure 5D**). Utilising both single colony WGS and post-enrichment metagenomics there were five K loci found only in ward surface samples most of these restricted to 1-2 samples, however KL123 was identified in four samples. There was one K loci found only in maternal stool (KL38), and no K loci found only in neonatal stool.

### Antimicrobial resistance (AMR) genes and virulence factors

There were 63 distinct antimicrobial resistance (AMR) genes detected in total across the single colony WGS isolates. Isolates had between 19-37 AMR genes (median of 29) per genome. There was variation in the number of AMR genes per isolate depending on the sample source (ANOVA; *p* = <0.0001), with isolates from the ward surfaces having the greatest mean number of AMR genes per isolate (mean = 30.1, SD = 3.3) and those from maternal stool having the lowest mean number of AMR genes per isolate (mean = 27.6, SD = 3.5).

At least one ESBL gene was found in every single colony WGS isolate (isolates were selected for 3GC resistance) (**Figure 6 & 7B**). ESBL genes of the *bla*_CTX-M-15_ type were found frequently (540/552 [97.8%]), whilst *bla*_CTX-M-14_ genes were less frequent (5/552 [0.9%]). TEM ESBL genes were also identified including *bla*_TEM-57_ (5/552 [0.9%]). SHV ESBL genes included, *bla*_SHV-28_ (172/552 [31.2%]) and *bla*_SHV-27_ (30/552 [5.4%]) (**Figure 6 & 7B**). The post-enrichment metagenomic samples that came from samples where an ESBL *K. pneumoniae* was identified were analysed for the presence of AMR genes, with the caveat that not all of these would necessarily be from *K. pneumoniae* (**Figure 1B**). There were 19 different *bla*_CTX-M_ genes identified of which the most frequently identified were *bla*_CTX-M-15_ (589/638 [92.3%]), *bla*_CTX-M-27_ (18/638 [2.8%]) and *bla*_CTX-M-14_ (11/638 [1.7%]; **Figure 7B**). TEM ESBL genes were also identified including *bla*_TEM-57_ (9/638 [1.4%]). There were 48 different SHV genes. SHV ESBL genes included, *bla*_SHV-28_ (153/638 [24.0%]) and *bla*_SHV-27_ (37/638 [5.8%]).

**Figure 6.**
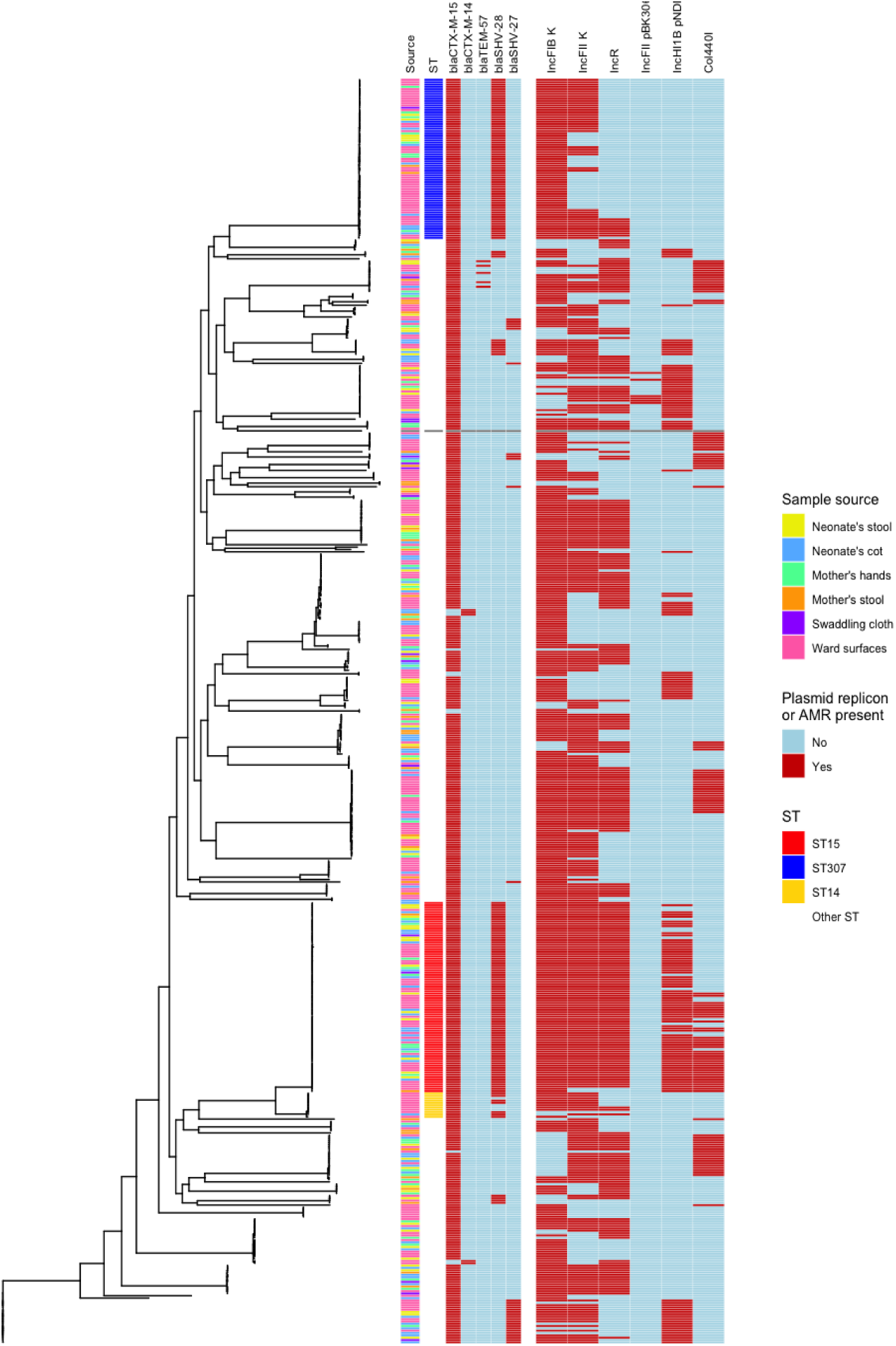
A phylogenetic tree of the ESBL *K. pneumoniae sensu strictu* isolates from the single colony isolates. The first column represents the sample source of the isolate, the second column represents the ST of the isolates, the third-fifth columns represent the presence or absence of specific AMR genes and the sixth-eleventh columns represent the presence or absence of specific plasmid replicons.

**Figure 7.**
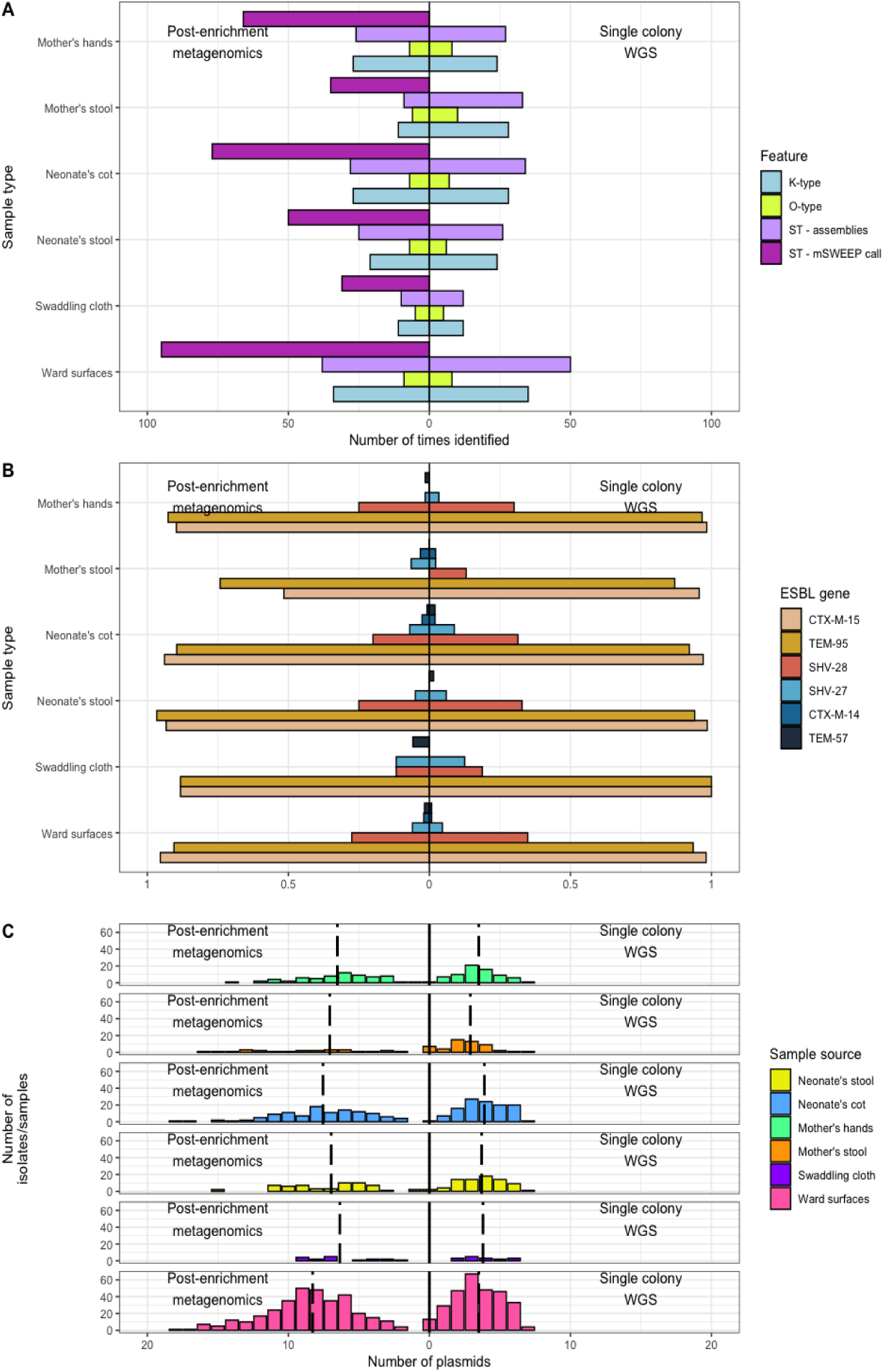
A) A bar chart showing the number of different genomic features (ST, O-type or K-type) that were identified in each sample type by post-enrichment metagenomic methods and single colony WGS. **B)** A bar chart showing the proportion of single colony WGS isolates or plate sweeps that contained different ESBL genes. **C)** A frequency chart showing the numbers of plasmids for *Klebsiella pneumoniae* isolate and post-enrichment metagenomic samples. The x axis represents the number of plasmids for each isolate and the y axis represents the number of isolates with that number of plasmids. The panels represent the different sample types and the vertical black dotted lines represent the mean number of plasmids for that sample type.

There were five single colony WGS isolates which had the *rmpA*, *iroB, salmochelin (iroD), aerobactin (iucA-D* and *iutA)* virulence factors. There was one cluster of three isolates of ST86 associated with one dyad and another cluster of isolates of ST25 associated with another dyad. All isolates encoded for the KL2 capsule type. ESBL producing isolates of ST86 with these same virulence determinants have been identified as causing invasive disease at the same site^5^. We also analysed the post-enrichment metagenomic data that came from samples where an ESBL *K. pneumoniae* was identified to investigate the presence of virulence factors. We excluded the *iroB* and *aerobactin (iucA-D* and *iutA)* virulence factors which occurred in between 77/624 (12.3%) and 132/624 (21.1%) samples depending on the specific virulence factor, in samples that contained 73 different *K. pneumoniae* STs, implying that they were most likely representing virulence factors from other bacterial species captured in the plate sweep. The most likely source of these virulence factors were *E. coli* (present in 123/132 [93.2%] samples), *Enterobacter cloacae* (present in 93/132 [70.5%] samples) or *Enterobacter asburiae* (present in 60/132 [45.5%] samples). *rmpA* was identified in eight samples, and *iroD* in nine samples (all but one of which also had *rmpA).* Three of these samples contained ST86 and six ST25. One sample was from ward surface swabs, and the others were all associated with the same two dyads that had single colony WGS isolates with these virulence factors.

### Plasmid replicons

The single colony WGS isolates had between 0 and 7 plasmid replicons, with a mean of 3.6 replicons per isolate and 22 replicons identified in total. There was variation in the mean number of plasmid replicons when categorized by sample type (ANOVA; *p* = 0.0086). Most sample types demonstrated mean plasmid numbers within the range of 3.5 to 3.9, however, isolates from mother’s stool exhibited a lower mean replicon number of 2.9 (**Figure 7C**). The predominant replicon identified was IncFIB K, followed by InFII K, IncR, FIA pBK30683, IncHI1B pNDM MAR, IncFIB pNDM Mar and Col440I. Additionally, two isolates from ST1799 and one from ST45, despite deriving from ESBL+ selective plates, has no known plasmid replicons identified, however the contigs containing *bla*_CTX-M-15_ on both isolates had high resemblance to *K. pneumoniae* plasmids in the NCBI database (**Supp. methods**) so were likely on plasmids not in the plasmidfinder database.

For the samples where ESBL^+^ *K. pneumoniae* was identified from the single-colony picks on selective plates, we also analyzed the post-enrichment metagenomic samples regarding plasmid replicons, recognizing the caveat that not all these plasmids will be associated with *K. pneumoniae.* We note between 0 and 20 plasmid replicons per sample, with a mean of 7.7 replicons identified in total (**Figure 7C**). The most frequently occurring replicons were like those in the single colony WGS, however there were 52 replicons which did not appear in the single colony WGS data, including 25 Inc type and 10 Col type. The number of replicons in the post-enrichment metagenomics also varied by sample type (ANOVA; *p* = 0.0002), however the pattern differed to that of the single colony WGS. Most sample types had between 6.4 and 7.1 replicons per sample (including maternal stool which had 7.1 replicons per sample, implying that despite having fewer plasmids associated with ESBL + *K. pneumoniae* the overall plasmid burden is similar in maternal stool and the other sample types) whilst cot swabs had 7.5 replicons per sample and surface swabs had 8.3 replicons per sample. Human associated samples had fewer replicons than environmental samples (6.8 vs 8.0; *p* = 0.0002; unpaired t-test).

## Discussion

This study assessed the diversity of *K. pneumoniae* carried by humans and present in the environment of a Malawian neonatal unit over a period of eight months, where *K. pneumoniae* neonatal sepsis is a leading cause of mortality. Understanding the true spectrum of pathogen diversity is crucial to assess potential reservoirs, differentiate the circulating diversity from high-risk hotspots and clones and ultimately to re-construct transmission networks. We used single-colony picks selecting for ceftriaxone resistance (ESBL-carrying isolates; a standard approach to evaluating ward level diversity) compared to limited-diversity metagenomics on matched plates without phenotypic selection, to assess within-sample diversity. We observed differences in ST distribution, O-type, AMR distribution and plasmid replicons by sample type. Post-enrichment metagenomics on non-ESBL selective media revealed greater ST diversity on the ward than was apparent through single colony WGS on ESBL selective media, whilst limiting the metagenomics data to strains with enough coverage resulting in assemblies largely reconstructed the single-colony pick diversity (**Figure 7A**).

In total, 121 different STs were identified, which is consistent with other studies that find high diversity of *K. pneumoniae* at single sites. A previous study examining a longitudinal collection of invasive isolates over a period of 20 years at this site found that ST15, ST14, ST25, ST1552, ST35 and ST39 frequently caused invasive infection and were all implicated in within-hospital transmission^5^. In our study the most frequently identified ST from single colonies was ST15, with marked prominence in dyad-associated samples, which may partly explain its impact as a frequent cause of human infection. ST15 has been implicated in the global dissemination of carbapenemase genes^48,49^ and is known as a successful hospital clone including at QECH, where it was a major ST as described in a large-scale analysis from 1996 to 2014^50^. ST307, which emerged in the 1990s^51^ and has now disseminated worldwide with outbreaks in South Africa^52^ and in Europe^53^ was also frequently identified in our study; and whilst less abundant than ST15 it was again frequently found in dyad-associated samples.

The most frequently detected ST from post-enrichment metagenomic data was ST14, which was found almost exclusively using this non-selective approach (indicating that many of these isolates were not ESBL-producers), and predominantly from ward surface swabs. A study of a *K. pneumoniae* outbreak on the Chatinkha nursery from 2014 indicated that ST340, ST14 and ST372 were dominant in that period, although only ST340 and ST372 seemed to be driving the outbreak, with ST14 present at similar frequency across the study duration^4^. Another study at this site confirmed ST14 as a frequent cause of infection across all ages^8^. This would be consistent with ST14 being present in the hospital environment but only occasionally acquiring resistance and causing infection, with other resistant clones persisting less in the environment. ST340 was also frequently found in our post-enrichment metagenomic samples and was more likely to be found in the ward surface swabs than human associated ones.

The apparent total ST diversity of *K. pneumoniae* in maternal stool was higher than for the rest of the samples, reflecting a higher between-sample diversity. Maternal stool samples were also less likely to have within-sample diversity, and the isolates had fewer AMR genes and plasmid replicons than ward surface swabs. This is likely because mothers are from diverse community locations and have thus been exposed to a wide range of *K. pneumoniae* they might acquire as commensal members of their gastrointestinal tract. Their microbiomes have also been exposed to less selective pressure in the form of antimicrobials. In contrast, the ward environment or neonate-associated (stool, cot, swaddling cloth) samples are linked by direct contact within a small area where colonizing bacteria are under selection pressure from frequent antimicrobial and antiseptic usage, elevated temperature, and high patient, guardian and healthcare worker footfall leading to a lot of potential transmission events. The neonatal ST diversity largely reflected that of cots and a subset of the diversity on ward surface swabs, as there is likely easy movement of bacteria between neonates and the ward environment, particularly the cots, which at the time of the study were wooden and very difficult to decontaminate adequately.

Whilst the O- and K-type diversity, like the STs, largely reflects similar patterns between the different sequencing approaches, notable differences can be observed with more diversity observed using single colony WGS than post-enrichment metagenomics. This could reflect the fact that only STs with enough coverage for assembly could have O- and K-types evaluated as these loci need assembling to identify the type. Greater identification of capsular polysaccharides from post-enrichment metagenomics approaches is likely with greater sequencing depth. The collectively greater diversity of mothers’ stool, also shows the most prominent differences in O- and K-type abundance between the two methods, which could be indicative of the same ST encoding different O- and/or K-types, as is often observed in *K. pneumoniae*. Despite some apparent variation however, the K- and O-type distribution are remarkably similar between the different sample types, again highlighting the high level of circulation between different compartments.

The post-enrichment metagenomic approach revealed extensive diversity at both the ward-level and within-sample level. For studies that are concerned with within-sample diversity (transmission studies and epidemiological descriptions), approaches such as post-enrichment metagenomics should be considered and methods will hopefully become further streamlined to allow efficient within-hospital tracking for real-time IPC investigations^21^. There was a large range of STs in individual samples ranging from one up to 19. This has been shown previously in *E. coli* but has not been as well-characterised for *K. pneumoniae*^14^. Previously this diversity might have been examined by taking multiple colony picks, which would have been unnecessary for most samples in this study and inadequate for a handful, or extremely deep (and thus costly) sequencing of samples directly.

A greater number of STs were found throughout the ward in the post-enrichment metagenomic samples compared to the single colonies. Some of these likely represent the population of non-ESBL *K. pneumoniae* that are selected against when ESBL selective media is used, whilst others represent the added diversity that is revealed when using post-enrichment metagenomics, and some may be spurious calls by the mSWEEP method. The detection of frequent co-occurrences of STs in the metagenomics data gives first indications of potential interactions between specific clones to be investigated further. In addition, identification of *K. pneumoniae* from non-selective sweeps in samples where no *K. pneumoniae* was isolated on selective agar strongly indicates that despite the high pressure of disinfectants, broadly antimicrobial sensitive isolates are present.

Read-based plasmid replicon detection shows a much larger diversity in samples from post-enrichment metagenomics (**Figure 7B**). However, it is important to note that plasmid replicons, AMR and to some extent also virulence genes are readily shared between species, whereas K- and O-types are almost exclusively specific for *K. pneumoniae* and thus might give a better assessment of the species diversity, as mobile elements likely represent not only the larger number of different *K. pneumoniae* strains in the sample, but also other bacteria carrying plasmids, in particular close relatives like *E. coli*, *Enterobacter* or the also often hospital-associated *Acinetobacter*.

Assessing the insights gained by our two approaches, whilst plate sweep metagenomics is an essential step towards an accurate understanding of the *K. pneumoniae* population, and to be able to infer transmission events, we note existing challenges that will need further exploration. Some key epidemiological markers of *K. pneumoniae* diversity (ST, K- and O-types) were under-identified using the post-enrichment metagenomics pseudo-assemblies compared to assembly-free ST assessment of the same sequence data using mSWEEP, which is likely due to an inability to assemble organisms found at low relative abundance in the samples. This may be improved in subsequent studies by increasing read depth. As the single colony WGS was performed on ESBL selective media and the post-enrichment metagenomics was not, firm conclusions about the differences between the two approaches are also difficult to draw.

In conclusion there was high diversity of *K. pneumoniae* on the ward, clear differences between hospital-related samples showing a close interaction network distinct from maternal stool, and notable within-sample diversity. We observed differences between major STs, with ST15, ST307 and ST14 all frequent in our study, but ST14 was only present in high numbers with the post-enrichment metagenomics. Further work on sample preparation and sequencing as well as best analysis strategy will be essential to implement plate sweeps routinely but indicates a significant improvement for future transmission studies. Our analysis further highlights the extensive spread of *K. pneumoniae* across the hospital environment and will allow detailed insights for focus of IPC efforts in this high-risk setting.

## Author contributions

The study was conceived by RL, NAF, JC, and CJ. Methodology was developed by NAF, CJ, RL, PM, TE, EH, PS, JC and OP. Software was developed by OP. Validation was performed by OP and AZ. Formal analysis was performed by OP, EH and CJ. Investigation was performed by HM, OP, PS and AZ. Resources were provided by NAF, NRT and CJ. Data curation was performed by OP. Writing the original draft was performed by OP, NAF, CJ, JC and EH. Reviewing and editing the manuscript was performed by all authors. Visualisation was performed by OP and EH. Supervision was performed by NAF, KK, ET, CJ, JC, EH and NRT. Project administration was performed by OP, RL and HM. Funding acquisition was performed by NAF and NRT. All authors read and approved the final manuscript.

## Data Availability

All sequencing data used in this article is freely available on European Nucleotide archive under project IDs PRJEB42462 (https://www.ebi.ac.uk/ena/browser/view/PRJEB42462), PRJEB40384 (https://www.ebi.ac.uk/ena/browser/view/PRJEB40384) and NEOTRACK project ID PRJEB63570 (https://www.ebi.ac.uk/ena/browser/view/PRJEB63570). Accession numbers are in Supplementary Table S1 along with relevant metadata. Details of bioinformatics analysis outputs on single colony WGS isolates are in Supplementary Table S2. Bioinformatics analysis outputs performed on plate sweeps are in Supplementary Table S3, the ST calls from mSWEEP are in Supplementary Table S4 and the bioinformatic outputs performed on the pseudo-assemblies generated by mGEMS are in Supplementary Table S5. A customised reference database used for mSWEEP and mGEMS is contained in the plaintext file mSWEEP_database_pearse.fasta, and the reference groups are in Supplementary Table S6. Details of the number of samples, isolates and plate sweeps are in Supplementary Table S7. Details of the environmental swabbing locations are in Supplementary Table S8. Details of PCR primers are in Supplementary Table S9. All code and required input files to construct the figures in this manuscript is provided at Github. The authors confirm all supporting data, code and protocols have been provided within the article or through supplementary data files.

https://www.ebi.ac.uk/ena/browser/view/PRJEB40384

https://www.ebi.ac.uk/ena/browser/view/PRJEB63570

## Acknowledgements

We would like to thank the clinical team on the Chatinkha nursery for working with us over the course of the study and beyond, we really enjoyed working with you and learnt a lot. The nursing team on the study who did a great job in recruiting patients. We would also like to thank the laboratory team in the Bacterial and Drug Resistant Infections (BDRI) group who helped us with sample processing and the pathogen informatics team at Wellcome Sanger Institute for assistance with bioinformatic analysis. We would also like to thank the team at Lancaster, particularly Barry Rowlingson, who helped us to generate patient wristbands. We would also like to thank the data team at MLW.

## Supplementary methods Study site

QECH is the government run tertiary referral centre for the Southern region of Malawi, which provides healthcare free at the point of delivery. It also serves the urban and rural population of Blantyre at primary and secondary level of care. Maternity care provided is comprehensive and 30% of births are by caesarean section^54^. There is a 3.4% stillbirth rate^55^. Chatinkha nursery is the neonatal unit at QECH, and of approximately 14,000 neonates born at QECH each year, 5,000 are admitted to Chatinkha nursery. Approximately 80% are born inside and 20% born outside QECH, predominantly in other healthcare facilities. It is extremely uncommon for a neonate to be admitted to Chatinkha nursery after discharge into the community. There are between 30 and 70 neonates on the ward at any one time. Any neonate born at QECH with problems during or after birth is admitted for management or observation, as well as neonates with complex medical or surgical needs from the district hospitals. Neonatal mortality from sepsis on the unit was 9% in 2021^56^. Neonates can receive oxygen therapy, continuous positive airway pressure, intravenous medication and nasogastric or oro-gastric tube feeding, but Chatinkha does not have neonatal intensive care facilities.

## Ward surface swabbing

Ward surface swabs, hand swabs and cot swabs were taken using a sponge-stick in 10ml of neutralising buffer (3M, UK). Oxygen tubes have a narrow diameter and were swabbed using a small rayon swab in neutralising buffer, attempting to cover the inside surface as well as the outside. Using aseptic non-touch technique, the swab was wiped over the relevant surface. We aimed to cover as much surface area as possible so for small items of equipment we swabbed the whole surface, for medium items of equipment, all high contact surfaces. For large areas such as floors we swabbed a random and representative area of the entire surface. For staff members, each week a single pooled hand swab was taken from at least 5 staff members.

## PCR details

Pink colonies on chromogenic agar were assumed to be *Escherichia coli*. White colonies were assumed *Acinetobacter baumannii* unless they had the morphological appearance of *Salmonella* spp. (flat and slightly colourless); these isolates underwent PCR to confirm *Salmonella* spp. Each single blue colony was picked off the plate and DNA was extracted by heating to 95°C in molecular grade water for 5 minutes, after which the extracted DNA underwent a multiplex High Resolution Melt PCR assay using primers specific to *K. pneumoniae*^20^ and *Enterobacter* spp., using Type-it HRM PCR Kit (Qiagen, Germany) according to the manufacturer’s instructions, with 400nM of each primer. Amplification was carried out using an QuantStudio 7 Flex Real-Time PCR System thermocycler (Applied Biosystems, USA) with a thermal profile of a hold step (95°C for 5 minutes), 30 PCR cycles (95°C for 10 seconds, followed by 58°C for 30 seconds and then 72°C for 20 seconds) followed by a melt curve step (75°C for 30 seconds then melting up to 95°C, with reads every 0.1°C). A post amplification HRM step was used to identify amplicon melt temperatures and assign the species ID.

## Single colony WGS assembly

We assembled isolates using SPAdes v3.10.0^57^, trialing different kmer lengths between 41 and 127 to find the optimal kmer length. An assembly improvement step was applied to the assembly with the best N50, contigs scaffolded using SSPACE v2.0^58^ and sequence gaps filled using GapFiller v1.11^59^. PROKKA v1.5^60^ with genus specific databases from RefSeq^61^ was used for annotation. The improved assembly step uses software developed by the Pathogen Informatics team at the WSI^62,63^.

## mSWEEP database creation

The database was based on the one used in the original mSWEEP publication^18^. The *Campylobacter jejuni* and *Staphylococcus epidermidis* genomes were removed, leaving the *K. pneumoniae* database and *E. coli* database plus individual examples of other bacterial species. To these we added 462 *K. pneumoniae* single colony WGS isolates analyzed in this study, as well as a collection of 429 *E. coli* genomes from Malawi/QECH (**Supp. Table 6**). The entire database included 1835 *K. pneumoniae*, 1940 *E. coli* and 26 other organisms. The labelling for the mSWEEP database included clonal complexes and STs, so all sample labels were converted to multi-locus sequence type.

## Quality control

Following single colony whole genome sequencing and subsequent quality control, 28 isolates were excluded, five due to between-species contamination, 27 due to failed assembly or poor assembly metrics and one due to within-species contamination with some isolates excluded for more than one reason (**Supp.** Figures 1, 2 and 3; **Figure 2A**), leaving 597 samples. In the case of multiple colony phenotypes resulting in more than one pick per plate, duplicated STs were removed, leaving 552 isolates in total.

772 (**Supp. Table 1 & 7**) plate sweeps from MacConkey agar were selected for post-enrichment metagenomic sequencing. We included samples that were positive for ESBL; this included 350 samples where only ESBL^+^ *K. pneumoniae* were grown from the same sample, and 188 from samples that were positive for ESBL^+^ *E. coli* but not *K. pneumoniae*. These were included to assess the *K. pneumoniae* diversity in the potential absence of ESBL^+^ *K. pneumoniae* strains. Finally, 216 samples were positive for both ESBL^+^ *E. coli* and *K. pneumoniae* and were also included, and 18 samples with missing laboratory metadata. All 772 metagenomic samples were used for diversity estimation in the samples (mSWEEP). Of these, 638 resulted in a prediction of at least one *K. pneumoniae* ST using mSWEEP. This includes 127 plate sweeps deriving from samples that had no *K. pneumoniae* growth on the ESBL^+^ selection plates samples, thus likely representing sensitive *K. pneumoniae* lineages in that sample (**Figure 1**).

## Subspecies identification from mSWEEP ST calls

Subspecies were identified from the mSWEEP ST calls using the supplementary data in this paper^47^. For STs not present in this dataset, they were identified using the BigsDB Institut Pasteur database, utilizing Kleborate v3.0.0^25^ on representative isolates from the database if the subspecies was not documented on the database.

## Plasmid identification

To determine the presence of plasmids in isolates that did not have plasmid replicons in the plasmidfinder database, contigs containing *bla*_CTX-M-15_ genes were run against the NCBI nucleotide database. They matched multiple sequences from plasmids, primarily in *K. pneumoniae*, with the closest match for all three being Klebsiella pneumoniae strain 6 plasmid pK006_3 (accession: CP034319.1; E value 0.0; accession length 143593).

## Data summary

All sequencing data used in this article is freely available on European Nucleotide archive under project IDs PRJEB42462 (https://www.ebi.ac.uk/ena/browser/view/PRJEB42462), PRJEB40384 (https://www.ebi.ac.uk/ena/browser/view/PRJEB40384) and NEOTRACK project ID PRJEB63570 (https://www.ebi.ac.uk/ena/browser/view/PRJEB63570). Accession numbers are in Supplementary Table S1 along with relevant metadata. Details of bioinformatics analysis outputs on single colony WGS isolates are in Supplementary Table S2. Bioinformatics analysis outputs performed on plate sweeps are in Supplementary Table S3, the ST calls from mSWEEP are in Supplementary Table S4 and the bioinformatic outputs performed on the pseudo- assemblies generated by mGEMS are in Supplementary Table S5. A customised reference database used for mSWEEP and mGEMS is contained in the plaintext file mSWEEP_database_pearse.fasta, and the reference groups are in Supplementary Table S6. Details of the number of samples, isolates and plate sweeps are in Supplementary Table S7. Details of the environmental swabbing locations are in Supplementary Table S8. Details of PCR primers are in Supplementary Table S9. All code and required input files to construct the figures in this manuscript is provided at Github. The authors confirm all supporting data, code and protocols have been provided within the article or through supplementary data files.

## Supplementary Figures

**Figure S1.**
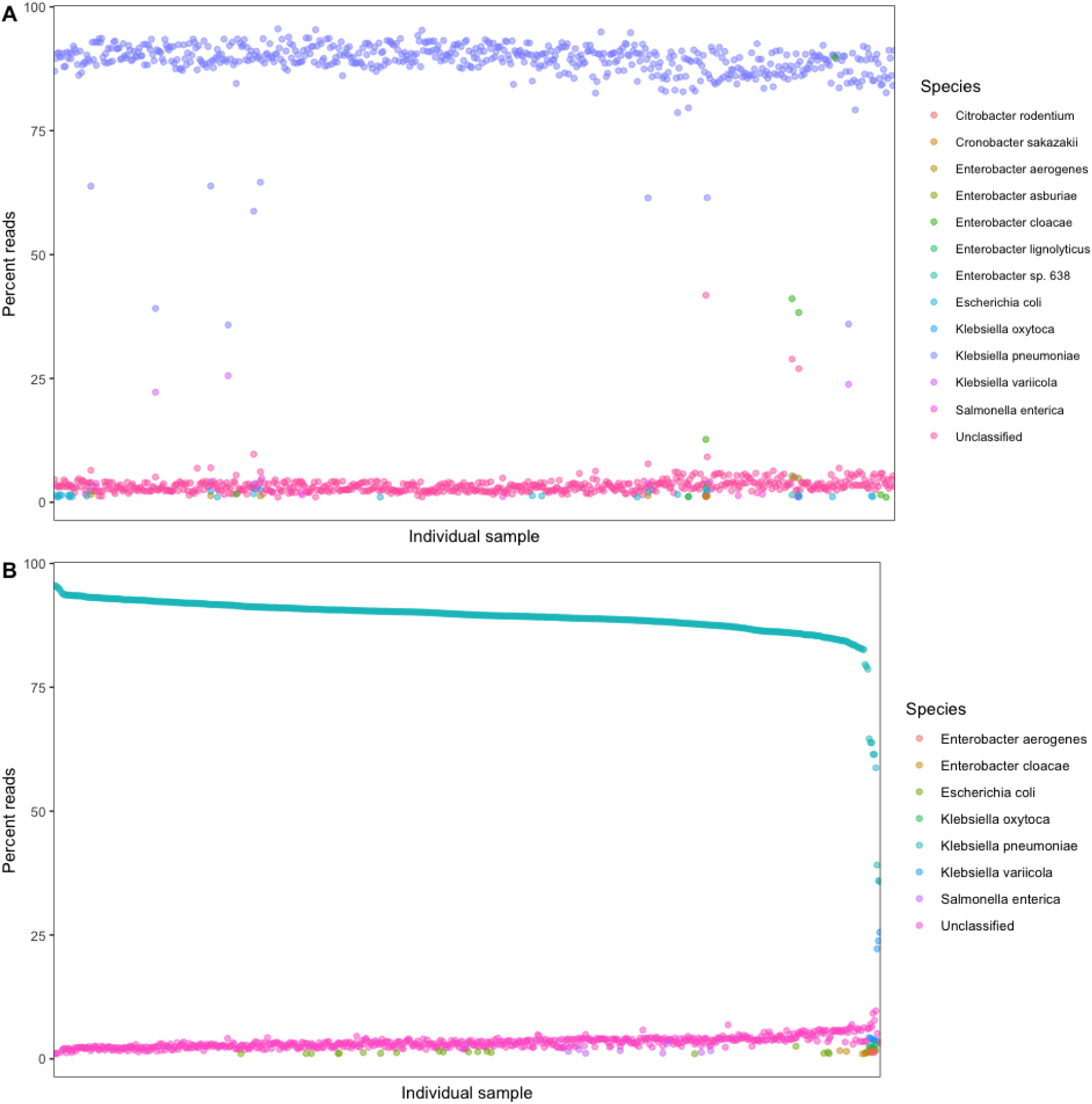
A) For all single colony WGS isolates; the percent of all sample reads assigned to different organisms. **B)** For single colony WGS isolates, with isolates containing greater than 5% reads assigned to an organism other than *K. pneumoniae*; the percent of sample reads with assigned to different organisms.

**Figure S2.**
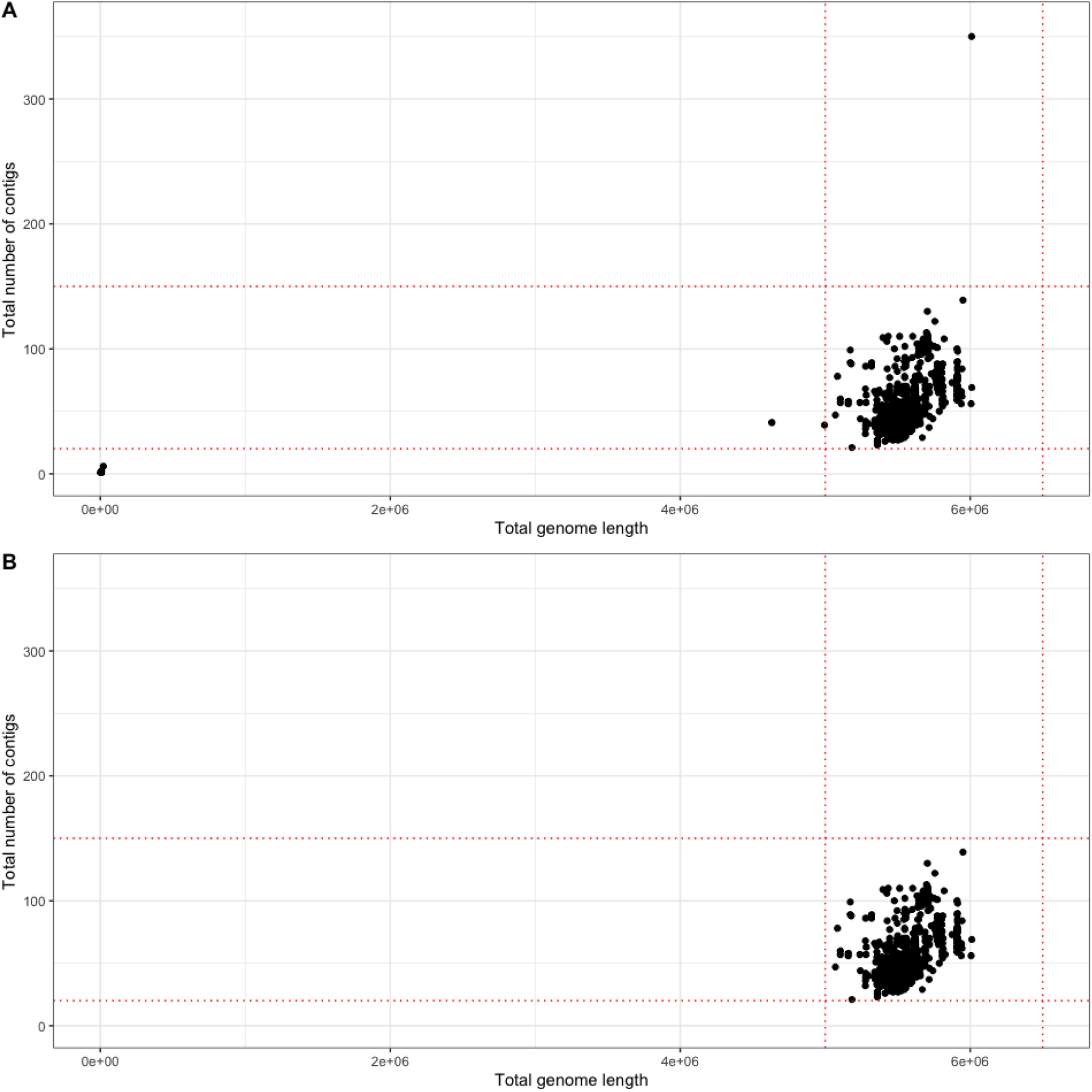
Scatter graphs showing the total genome length against the total number of contigs for each single colony WGS isolate, dotted red lines show the QC thresholds. **A)** Pre-QC. **B)** Post-QC.

**Figure S3.**
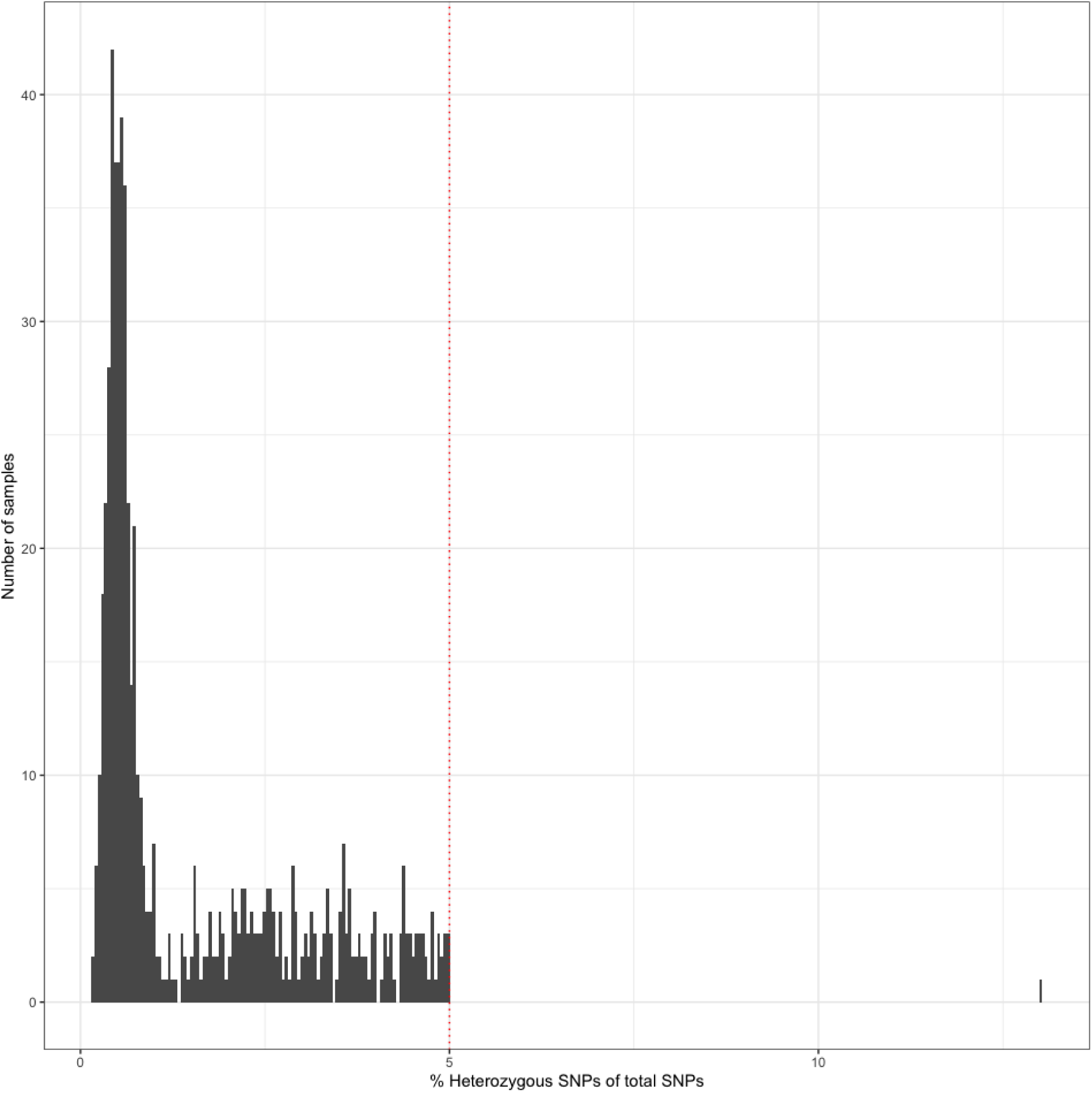
A histogram showing the frequency of single colony WGS isolates with numbers of heterozygous SNPs, the red dotted line represents the cut off of 5%, above which isolates were excluded.

